# CCR5 deficiency: decreased neuronal resilience to oxidative stress and increased risk of vascular dementia

**DOI:** 10.1101/2023.02.15.23285967

**Authors:** Benjamin B. Tournier, Silvia Sorce, Antoine Marteyn, Roberta Ghidoni, Luisa Benussi, Giuliano Binetti, François R Herrmann, Karl-Heinz Krause, Dina Zekry

**Author notes:** **Corresponding author:** Dr Benjamin B. Tournier, PhD, Division of Adult Psychiatry, Department of Psychiatry, Geneva University Hospitals, Avenue de la Roseraie, 64, 1205 Geneva, Switzerland. DZ and K-HK contributed equally to this work.

## Abstract

**Introduction:** As the chemokine receptor5 (CCR5) may play a role in ischemia, we studied the links between CCR5 deficiency, the sensitivity of neurons to oxidative stress, and the development of dementia.

**Methods:** Logistic regression models with CCR5 and ApoE polymorphisms were applied to 205 cognitively normal and 189 dementia patients included in Geneva. The impact of oxidative stress on *Ccr5* expression and cell death was assessed in mice neurons.

**Results:** CCR5-Δ32 allele synergized with ApoEε4 as risk factor for dementia and specifically for dementia with a vascular component. We confirmed these results in an independent cohort from Italy (157 cognitively normal and 620 dementia). Carriers of the ApoEε4/CCR5-Δ32 genotype aged ≥80 years have an eleven-fold greater risk of vascular and mixed dementia. Oxidative stress induced cell death in *Ccr5*^*-/-*^ mice neurons.

**Discussion:** We propose the vulnerability of CCR5-deficient neurons in response to oxidative stress as possible mechanisms contributing to dementia.

## Introduction

The chemokine receptor 5 (CCR5) is a G protein-coupled receptor mainly expressed in immune cells, where, after stimulation by its specific ligands (chemokine ligand 3 -CCL3-, CCL4 and CCL5), it regulates chemotaxis and cell activation [1]. Within the central nervous system, CCR5 is expressed by glial cells, neurons as well as endothelial and vascular smooth muscle cells. CCR5 is involved in regulating the inflammatory response, learning and memory processes as well as pathological cell death [2]. Although it has been demonstrated that both CCR5 and its ligands are upregulated in some pathological situations including Alzheimer’s disease (AD) [2], the beneficial impact of CCR5 expression on cognitive outcome of mouse AD model is still controverted [3-5]. In humans, a 32-base-pair deletion is responsible to the occurrence of a premature stop codon into the CCR5 receptor locus (CCR5-Δ32), which lead to natural receptor dysfunction. Surprisingly, no link between the CCR5 deletion and the risk of developing AD were revealed [5, 6]. In very old individuals, pure AD and pure vascular dementia are frequent, but mixed dementia is even more prevalent [7]. While the rare cases of AD in young patients are monogenetic, the risk for old-age dementia is thought to be modulated by a large number of genetic variants with relatively low penetrance, but high prevalence [8]. Until now, ApoEε4 is the only known genetic risk factor strongly associated with old age dementia [9]. Also, hitherto no specific risk factors for old age AD versus vascular or mixed dementia have been identified. The implication of CCR5-Δ32 in the development of vascular dementias and mixed dementias is not known. As ApoEε4 represents a risk factor for both AD and for vascular dementias, we hypothesize that the CCR5-Δ32/ApoEε4 polymorphism combination could therefore represent a greater risk factor than the two polymorphisms taken separately.

Oxidative stress induced by the release of reactive oxygen species (ROS) is a key player of neuronal death and neurodegenerative diseases [10-12]. In vascular disorders, as ROS/oxidative stress are increased, it can be suspected that they participate to neuronal death and dementia [13]. ROS promote the activation of specific transcription factors, such as p53 or NF-κB, which control the expression of death/survival-related genes [14]. Neuronal death is increased in response to nerve transaction, and brain damage and neuronal death are increased after cerebral stroke in *Ccr5*^*-/-*^ deficient mice as compared to control [15, 16]. Thus, these preliminary studies supposed a role of CCR5 in neuroprotective mechanisms [17]. However, the factors regulating CCR5 up-regulation in neurons have not been elucidated. Thus, the knowledge of the functional role of CCR5 in oxidative stress environment is lacking.

The aims of the present study were to analyze the impact of CCR5 deletion on (i) the risk of dementia in very old subjects, and on (ii) the molecular response of neurons to oxidative stress.

## Results

394 subjects were enrolled in Geneva. Two of the 394 subjects were homozygous for the CCR5-Δ32 allele (CCR5-Δ32/CCR5-Δ32: 0.5%), 86 were heterozygous (CCR5^+^/CCR5-Δ32: 21.83%), and 306 were homozygous for the wild-type (CCR5^+^/CCR5^+^: 77.67·%). 205 were cognitively normal, and 189 were demented (73 AD, 20 vascular dementia, 82 with mixed dementia and 14 with other dementias). The frequency of the CCR5-Δ32 allele is not different between the groups (Chi^2^, p = 0.18). The frequency of the ApoEε4 allele is lower in cognitively normal (12.2%) compared to patients with dementia of various aetiologies (26.5%; Chi^2^, p = 0.0003). Age and gender did not differ between the groups (mean age: 85.1 ± 6.8; Chi^2^, p = 0.120; women: 76 %; Chi^2^, p =0.345). Table 1 summarises the demographics, CCR5-Δ32 and ApoEε4 allele frequencies as a function of cognitive diagnosis. To measure the impact of cerebral vascular lesions, patients with vascular and mixed dementia were analysed together.

**Table 1.**
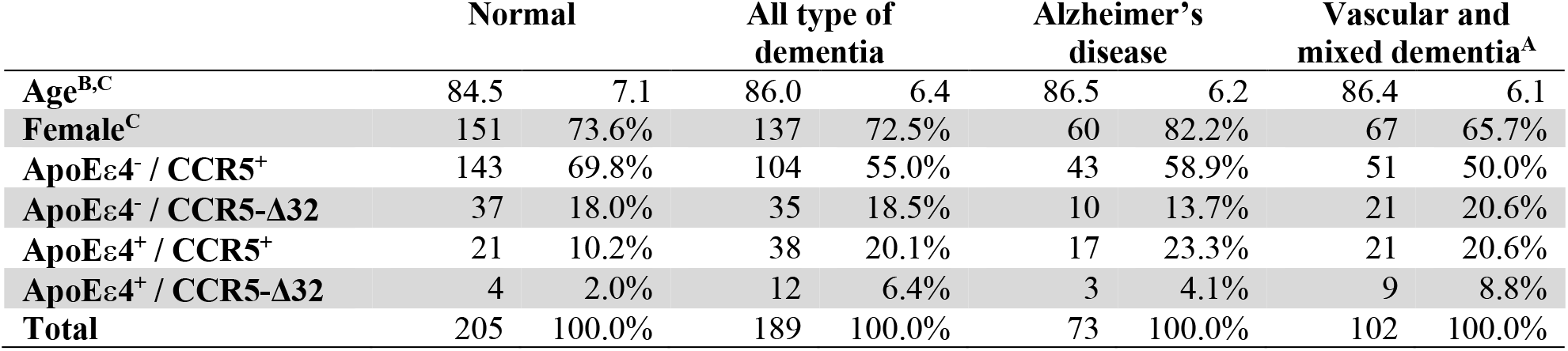
Comparison of CCR5-Δ32 and ApoEε4 allele frequencies between dementia patients, including the various dementia aetiologies, and subjects that are cognitively normal in the Swiss population. Data are expressed as number of cases and %. ^A^Patients with vascular and mixed dementia were pooled to measure the impact of cerebral vascular lesions. ^B^Data are expressed as means ± SD. ^C^There is no statistically difference between cognitively normal subjects and patients with dementia of various aetiologies (Chi^2^; p = 0.120 for age and p = 0.345 for sex ratio). ApoEε4^**+**^: one or two copies of ApoEε4; ApoEε4^**-**^: no copies of ApoEε4; CCR5-Δ32: one or two copies of the CCR5-Δ32 bp deleted allele; CCR5^**+**^: two copies of the CCR5 wild-type allele.

### Univariate and multiple logistic regression analysis

Table 2 shows for the Swiss sample univariate and multiple logistic regression analyses, adjusted for age and sex, including the predictive variables tested: presence or absence of dementia, and dementia aetiology (AD or dementia with a vascular component [vascular and mixed dementia grouped]).

**Table 2.** Univariate and multiple logistic regression models of the risk of dementia and of its main aetiologies with cognitively normal subjects as the reference group (n = 205) according to CCR5 and ApoE genotypes in the Swiss population. ^A^ApoEε4^+^: one or two copies of ApoEε4; ApoEε4^-^: no copies of ApoEε4; ^B^CCR5-Δ32: one or two copies of the CCR5-Δ32 bp deleted allele; CCR5^+^: two copies of the CCR5 wild-type allele. ^**C**^Bold entries = relevant results.

|  | *Dementia*<br>(n = 189) |  |  | *Alzheimer's disease*<br>(n = 73) |  |  | *Vascular or mixed dementia*<br>(n = 102) |  |  |
| --- | --- | --- | --- | --- | --- | --- | --- | --- | --- |
| Univariate logistic regression |  |  |  |  |  |  |  |  |  |
|  | Crude OR | 95% CI | p | Crude OR | 95% CI | p | Crude OR | 95% CI | p |
| ApoEε4<sup>+A</sup> | 2.59 | 1.52-4.39 | <0.001 | **2.72<sup>C</sup>** | **1.40-5.27** | **0.003** | **3.00** | **1.65-5.45** | **<0.001** |
| CCR5-Δ32<sup>B</sup> | 1.32 | 0.82-2.13 | 0.247 | 0.87 | 0.43-1.73 | 0.685 | 1.67 | 0.97-2.88 | 0.067 |
| Age | **1.04** | **1.00-1.07** | **0.018** | 1.05 | 1.01-1.09 | 0.031 | **1.04** | **1.01-1.08** | **0.021** |
| Male vs female | 1.06 | 0.68-1.65 | 0.790 | 0.61 | 0.31-1.19 | 0.146 | 1.46 | 0.87-2.44 | 0.148 |
| Multiple logistic regression |  |  |  |  |  |  |  |  |  |
|  | Adjusted OR | 95% CI | p | Adjusted OR | 95% CI | p | Adjusted OR | 95% CI | p |
| *Association* |  |  |  |  |  |  |  |  |  |
| *ApoEε4 / CCR5Δ32* |  |  | **0.002** |  |  | **0.026** |  |  | **<0.001** |
| ApoEε4<sup>-</sup> / CCR5<sup>+</sup> | 1.00 | -- | -- | 1.00 | -- | -- | 1.00 | -- | -- |
| ApoEε4<sup>-</sup> / CCR5-Δ32 | 1.30 | 0.77-2.20 | 0.328 | 0.90 | 0.41-1.96 | 0.788 | 1.59 | 0.85-2.97 | 0.144 |
| ApoEε4<sup>+</sup> / CCR5<sup>+</sup> | **2.49** | **1.38-4.49** | **0.002** | **2.69** | **1.30-5.56** | **0.007** | **2.80** | **1.41-5.56** | **0.003** |
| ApoEε4<sup>+</sup> / CCR5-Δ32 | **4.13** | **1.29-13.1** | **0.017** | 2.49 | 0.54-11.58 | 0.243 | **6.31** | **1.86-21.3** | **0.003** |
| *Adjusted for age and sex* |  |  |  |  |  |  |  |  |  |
|  |  |  | **<0.001** |  |  | **0.005** |  |  | **<0.001** |
| ApoEε4<sup>-</sup> / CCR5<sup>+</sup> | 1.00 | -- | -- | 1.00 | -- | -- | 1.00 | -- | -- |
| ApoEε4<sup>-</sup> / CCR5Δ32 | 1.35 | 0.79-2.30 | 0.265 | 0.91 | 0.41-1.99 | 0.814 | 1.70 | 0.90-3.22 | 0.100 |
| ApoEε4<sup>+</sup> / CCR5<sup>+</sup> | **2.47** | **1.36-4.47** | **0.003** | **2.80** | **1.34-5.86** | **0.006** | **2.81** | **1.40-5.63** | **0.004** |
| ApoEε4<sup>+</sup> / CCR5Δ32 | **4.46** | **1.39-14.4** | **0.012** | 3.57 | 0.72-17.78 | 0.121 | **7.27** | **2.09-25.2** | **0.002** |
| Age | **1.04** | **1.00-1.07** | **0.018** | **1.05** | **1.01-1.10** | **0.026** | 1.04 | 1.01-1.09 | 0.015 |
| Male vs female | 1.12 | 0.71-1.78 | 0.612 | 0.53 | 0.26-1.07 | 0.078 | 1.61 | 0.94-2.75 | 0.082 |

### Dementia

In univariate analysis, the ApoEε4 allele was found to be an independent predictor of dementia (OR = 2.59, 95% CI = 1.52–4.39, p < 0.001) even after adjustment for age and sex; while it was not the case for the CCR5-Δ32 allele (OR = 1.32, 95% CI = 0.82–2.13, p = 0.247). Introducing all the variables into the model showed that the ApoEε4 allele remained statistically significant (OR = 2.49, 95% CI = 1.38–4.49). However, it presented a greater significance when associated with the CCR5-Δ32 allele, and the risk of dementia increased to four times that of cognitively normal patients (OR = 4.13, 95% CI = 1.29–13.15). After adjusting for age and sex, the model remained significant (p < 0.001), with the OR for ApoEε4 combined with CCR5-Δ32 allele-carrying genotypes reaching 4.46 (95% CI = 1.39–14.42) (Table 2).

### Alzheimer’s disease

In univariate analysis, the ApoEε4 allele was also found to be an independent predictor of AD (OR = 2.72, 95% CI = 1.40–5.27, p = 0.003) even after adjustment for age and sex; while it was not the case for the CCR5-Δ32 allele (OR = 0.87, 95% CI = 0.43–1.73). The introduction of all the variables into the model showed that the ApoEε4 allele was the only statistically significant predictor of AD, even after adjusting for age and sex (OR = 2.80, 95% CI = 1.34–5.86, p = 0.006).

### Vascular and Mixed dementia (dementia with a vascular component)

In univariate analysis, the ApoEε4 allele was found to be an independent predictor of the outcome (OR = 3.0, 95% CI = 1.65–5.45), while it was not the case for the CCR5-Δ32 allele which shows only a trend (OR = 1.67, 95% CI = 0.97–2.88, p = 0.067). The introduction of all the variables into the model showed that the ApoEε4 allele remained statistically significant (OR = 2.80, 95% CI = 1.41–5.56). However, when associated with the CCR5-Δ32 allele, the significance was greater than with ApoEε4 alone, and the risk of dementia increased up to six times (OR = 6.31, 95% CI = 1.86– 21.38). The model remained significant after adjusting for age and sex, and ApoEε4 combined with CCR5-Δ32 allele-carrying genotypes increased the risk of dementia by a factor of 7.27 (95% CI = 2.09–25.2, p = 0.002).

### Validation population

To validate our results, we investigated 777 consecutively enrolled subjects (mean age 78.6 ± 6.4; 69.8 % women) from a memory clinic in Brescia; 157 were cognitively normal, and 620 with dementia (319 AD; 125 vascular dementia, 176 with mixed dementia, Table 3). 7 of the 777 subjects were homozygous for the CCR5-Δ32 allele (CCR5-Δ32/CCR5-Δ32: 1.0%), 84 were heterozygous (CCR5^+^/CCR5-Δ32: 9.6%), and 686 were homozygous for the wild-type (CCR5^+^/CCR5^+^: 89.5%). Table 3 summarises the demographics, CCR5-Δ32 and ApoEε4 allele frequencies as a function of cognitive diagnosis. Table 4 shows the results for the Italian sample regarding univariate and multiple logistic regression analyses. The odds ratio predicting vascular and mixed dementia in the unadjusted model were 1.26 (95% CI = 0.65-2.43; p = 0.49) for CCR5-Δ32 alone, 3.42 (95% CI = 2.13-5.49; p < 0.001) ApoEε4 alone, and 4.92 (95% CI = 1.09-22.2; p = 0.038) for ApoEε4 combined with CCR5-Δ32. After adjusting for age and sex, the progression in the odds ratio follows the same pattern, but the ApoEε4 combined with CCR5-Δ32 was not significant (p = 0.076). The fact that in the age- and sex-adjusted model significance was not reached, is most likely due to the relatively low prevalence of CCR5-Δ32 heterozygotes in the Italian sample, but possibly also due to the younger age (see below). The odds ratio predicting AD was significant for ApoEε4 (3.71, 95% CI = 2.27-6.07; p < 0.001) and ApoEε4 combined with CCR5-Δ32 (4.98, 95% CI = 1.10-22.5; p = 0.037). After adjusting for age and sex, only ApoEε4 (4.28, 95% CI = 2.45-7.47; p < 0.001) was significantly associated with AD risk (ApoEε4^+^/CCR5-Δ32: 2.81, 95% CI = 0.56-14.1; p = 0.21). We therefore performed a pooled analysis of the two populations.

**Table 3.** Comparison of CCR5-Δ32 and ApoEε4 allele frequencies between dementia patients, including the main dementia aetiologies, and subjects that are cognitively normal in the Italian population. To measure the impact of cerebral vascular lesions, patients with vascular and mixed dementia were analysed together. Data are expressed as number of cases and %. ^A^Patients with vascular and mixed dementia were pooled to measure the impact of cerebral vascular lesions. ^B^Data are expressed as means ± SD. ^**C**^Bold entries = relevant results. ApoEε4^+^: one or two copies of ApoEε4; ApoEε4^-^: no copies of ApoEε4; CCR5-Δ32: one or two copies of the CCR5-Δ32 bp deleted allele; CCR5^+^: two copies of the CCR5 wild-type allele.

|  | Normal |  | All type of dementia |  | Alzheimer's disease |  | Vascular and mixed dementia<sup>A</sup> |  |
| --- | --- | --- | --- | --- | --- | --- | --- | --- |
| **Age<sup>B,C</sup>** | 75.0 | 4.8 | 79.6 | 6.4 | 80.8 | 5.7 | 78.3 | 7.0 |
| **Female<sup>C</sup>** | 87 | 55.4% | 455 | 73.4% | 246 | 77.1% | 209 | 69.4% |
| **ApoEε4<sup>-</sup> / CCR5<sup>+</sup>** | 118 | 75.2% | 310 | 50.0% | 154 | 48.3% | 156 | 51.8% |
| **ApoEε4<sup>-</sup> / CCR5-Δ32** | 12 | 7.6% | 51 | 8.2% | 31 | 9.7% | 20 | 6.6% |
| **ApoEε4<sup>+</sup> / CCR5<sup>+</sup>** | 25 | 15.9% | 233 | 37.6% | 121 | 37.9% | 112 | 37.2% |
| **ApoEε4<sup>+</sup> / CCR5-Δ32** | 2 | 1.3% | 26 | 4.2% | 13 | 4.1% | 13 | 4.3% |
| **Total** | 157 | 100.0% | 620 | 100.0% | 319 | 100.0% | 301 | 100.0% |

**Table 4.** Univariate and multiple logistic regression models of the risk of dementia and of its main aetiologies with cognitively normal subjects as the reference group (n = 157) according to CCR5 and ApoE genotypes in the Italian population. ^A^ApoEε4+: one or two copies of ApoEε4; ApoEε4-: no copies of ApoEε4; ^B^CCR5Δ32+: one or two copies of the CCR5-32 bp deleted allele; CCR5Δ32-: two copies of the CCR5 wild-type allele. ^**C**^Bold entries = relevant results.

|  | *Dementia*<br>(n = 620) |  |  | *Alzheimer's disease*<br>(n = 319) |  |  | *Vascular or mixed dementia*<br>(n = 301) |  |  |
| --- | --- | --- | --- | --- | --- | --- | --- | --- | --- |
| Univariate logistic regression |  |  |  |  |  |  |  |  |  |
|  | Crude OR | 95% CI | p | Crude OR | 95% CI | p | Crude OR | 95% CI | p |
| ApoEε4<sup>+A</sup> | 3.45 | 2.22-5.39 | <0.001 | 3.49<sup>C</sup> | 2.18-5.58 | <0.001 | 3.42 | 2.13-5.49 | <0.001 |
| CCR5-Δ32<sup>B</sup> | 1.45 | 0.80-2.64 | 0.225 | 1.63 | 0.87-3.08 | 0.129 | 1.26 | 0.65-2.43 | 0.494 |
| Age | 1.13 | 1.09-1.16 | <0.001 | 1.22 | 1.17-1.28 | <0.001 | 1.09 | 1.05-1.12 | <0.001 |
| Male vs female | 0.45 | 0.31-0.65 | <0.001 | 0.37 | 0.25-0.56 | <0.001 | 0.55 | 0.37-0.82 | 0.003 |
| Multiple logistic regression |  |  |  |  |  |  |  |  |  |
|  | Adjusted OR | 95% CI | p | Adjusted OR | 95% CI | p | Adjusted OR | 95% CI | p |
| *Association ApoEε4 / CCR5Δ32* |  |  |  |  |  |  |  |  |  |
| ApoEε4<sup>-</sup> / CCR5<sup>+</sup> | 1.00 | -- | -- | 1.00 | -- | -- | 1.00 | -- | -- |
| ApoEε4<sup>-</sup> / CCR5-Δ32 | 1.62 | 0.83-3.14 | 0.155 | 1.98 | 0.97-4.02 | 0.059 | 1.26 | 0.59-2.68 | 0.547 |
| ApoEε4<sup>+</sup> / CCR5<sup>+</sup> | 3.55 | 2.23-5.64 | <0.001 | 3.71 | 2.27-6.07 | <0.001 | 3.39 | 2.07-5.56 | <0.001 |
| ApoEε4<sup>+</sup> / CCR5-Δ32 | 4.95 | 1.16-21.1 | 0.031 | 4.98 | 1.10-22.5 | 0.037 | 4.92 | 1.09-22.2 | 0.038 |
| *Adjusted for age and sex* |  |  |  |  |  |  |  |  |  |
| ApoEε4<sup>-</sup> / CCR5<sup>+</sup> | 1.00 | -- | -- | 1.00 | -- | -- | 1.00 | -- | -- |
| ApoEε4<sup>-</sup> / CCR5Δ32 | 1.28 | 0.64-2.57 | 0.48 | 1.66 | 0.73-3.75 | 0.223 | 1.13 | 0.52-2.47 | 0.753 |
| ApoEε4<sup>+</sup> / CCR5<sup>+</sup> | 3.99 | 2.45-6.5 | <0.001 | 4.28 | 2.45-7.47 | <0.001 | 3.71 | 2.21-6.21 | <0.001 |
| ApoEε4<sup>+</sup> / CCR5Δ32 | 3.48 | 0.80-15.2 | 0.098 | 2.81 | 0.56-14.1 | 0.210 | 3.99 | 0.86-18.3 | 0.076 |
| Age | 1.12 | 1.08-1.16 | <0.001 | 1.21 | 1.16-1.27 | <0.001 | 1.08 | 1.05-1.12 | <0.001 |
| Male vs female | 0.49 | 0.33-0.73 | <0.001 | 0.40 | 0.25-0.66 | <0.001 | 0.55 | 0.36-0.85 | 0.007 |

### Pooled results and effect of age

After pooling the Swiss and Italian populations (n= 1171, Supplemental Table 2) and contrasting the control group (n= 362) with the vascular and mixed dementia (dementia with a vascular component) group (n= 403), the logistic regression model adjusted for age as a continuous variable, sex and country showed that the ApoEε4 allele was still statistically significant (OR = 3.21, 95% CI = 2.22-4.63; p < 0.001; Supplemental Table 3). When associated with the CCR5-Δ32 allele, the risk of dementia with vascular component increased to six times that of normal patients (OR = 5.94, 95% CI = 2.19-16.1; p < 0.001). When repeating the multiple logistic regression model in the 494 subjects below the age of 80, a trend towards a significant effect was observed when considering the odds ratio associated with ApoEε4 combined with CCR5-Δ32 (OR = 3.43, 95% CI = 0.95-12.4; p < 0.059, Supplemental table 4). Of note, considering the 677 subjects aged ≥80, although the impact of ApoEε4 by itself was significant (OR = 2.42, 95% CI =1.32-4.42; p = 0.004), the odds ratio associated with ApoEε4 combined with CCR5-Δ32 reached 11.19 (95% CI = 2.36-53.0; p = 0.002, Supplemental table 5). These results were confirmed by repeating the multiple logistic regression model in the 1171 subjects combining age and genotype status: ApoEε4 combined with CCR5-Δ32 in subjects <80 years was not significant (OR = 3.40, 95% CI = 0.94-12.3; p = 0.063), whereas it was very high in subjects ≥80 years (OR = 10.55, 95% CI = 2.21-50.2; p = 0.003). Considering AD risk, the logistic regression model adjusted for age as a continuous variable, sex and country showed that the ApoEε4 allele was statistically significant (OR = 3.7, 95% CI = 2.39-5.72; p < 0.001; Supplemental Table 3) and, when associated with the CCR5-Δ32 allele, the risk of AD increased to 4 times that of normal patients (OR = 4.03 95% CI = 1.15-14.1; p = 0.029; Supplemental Table 3). However, stratifying by age, the risk for AD was not increase neither in subjects <80 years nor in subjects ≥80 years, with and without combining age and genotype status (subjects <80 years: OR = 2.22, 95% CI = 0.44-11.14; p = 0.331 and OR = 2.36, 95% CI = 0.41-13.47; p = 0.334 and subjects ≥80 years: OR = 4.86, 95% CI = 0.88-26.8; p = 0.07 and OR = 6.11, 95% CI = 0.94-39.6; p = 0.058, with and without combining age and genotype status, respectively).

### Expression of Ccr5 in neurons is increased by ROS-dependent neurotoxic stimuli

To demonstrate possible links between the presence of vascular disorders leading to dementia and the absence of CCR5, a study of the reactivity of CCR5 deficient neurons was conducted. To mimic the consequences of a vascular troubles, cortical neurons were exposed to several neurotoxic stimuli. Thus, glucose deprivation (associated with cobalt chloride) and excitotoxic concentrations of glutamate were used to mimics a hypoxic environment [18] and H_2_O_2_ was used to directly induce oxidative stress. All treatments increased *Ccr5* mRNA levels in neurons from WT animals (Figure 1A). As hypoxia/glucose deprivation and glutamate exert their neurotoxic action at least in part through generation of reactive oxygen species (ROS) [19, 20], oxidative stress might be a common denominator of these stimuli that induces *Ccr5* up-regulation.

**Figure 1.**
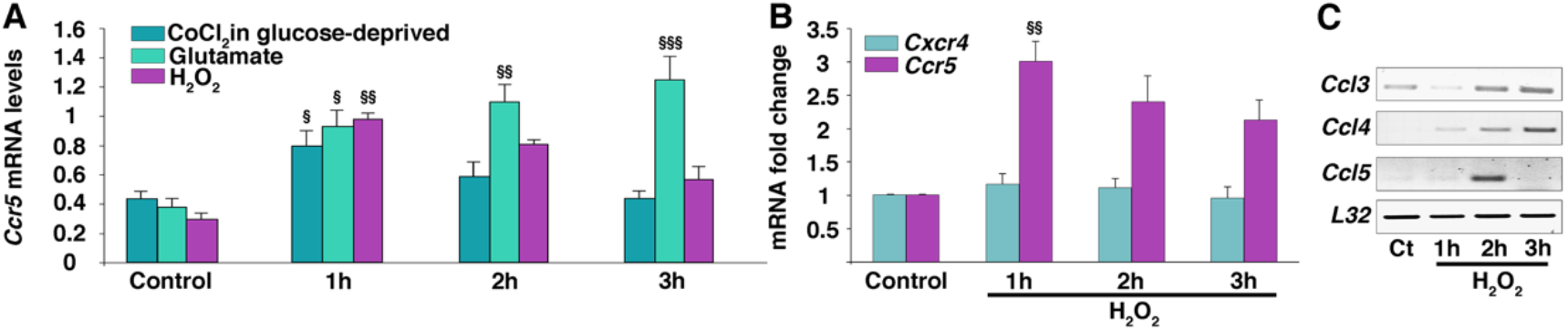
Oxidative stress increases Ccr5 expression and induces CCR5 ligands. **A**. Induction of *Ccr5* mRNA expression by oxidative stress in wild-type primary neurons. Wild-type primary neurons at day 10 *in vitro* were treated for the indicated time with CoCl_2_ (500 μM) in a glucose-deprived (GD) medium, glutamate (100 μM), H_2_O_2_ (30 μM) or vehicle (control, Ct). The levels of mRNA were determined by Real-Time PCR (*n*=4). **B**. The levels of *Ccr5* and *Cxcr4* mRNA were determined by Real-Time PCR.*\*\*p*<0.01 H_2_O_2_ 1h *vs*. ctr. **C**. Induction of CCR5 ligands (*Ccl3, Ccl4* and *Ccl5*) by H_2_O_2_ (30 μM). RT-PCR was performed by using the ribosomal *L32* house-keeping gene as control. Similar results were obtained from four independent experiments. ^§^*p*<0.05, ^§§^*p*<0.01 and ^§§§^*p*<0.001 as compared to the respective group control, using one-way ANOVA followed by Tukey *post-hoc* test.

Therefore, we investigated further the impact of H_2_O_2_-induced oxidative stress on *Ccr5* expression. A 3-fold above control increase in *Ccr5* mRNA within 1 hour of H_2_O_2_ exposure was shown with a slow decline in the next 2 hours (Figure 1B). The expression of *Cxcr4* (another chemokine receptor expressed in neurons [21]) was not affected by H_2_O_2_ indicating the specificity of oxidative stress-induced *Ccr5* mRNA (Figure 1B). Interestingly, mRNA levels of the CCR5 ligands *Ccl3, Ccl4* and *Ccl5* increased in response to H_2_O_2_ (Figure 1C), suggesting CCR5 receptor activation under our experimental conditions.

### Involvement of NF-κB in neuronal Ccr5 up-regulation

Next, we investigated potential mechanisms of ROS-dependent *Ccr5* up-regulation. By *in silico* analysis, we identified two putative binding sites for the NF-κB subunit p65/RelA within the regulatory regions of the murine *Ccr5* gene (Figure 2A). We then performed chromatin immunoprecipitation assay, in order to demonstrate the binding of NF-κB to *Ccr5* regulatory regions. After treatment of neurons with H_2_O_2_ or vehicle, chromatin was extracted and immunoprecipitated with the NF-κB antibody. We revealed by PCR that the transcription factor was binding to the two regulatory sequences of the *Ccr5* promoter in the H_2_O_2_-treated sample, while no bands were present in the control. Also, by performing PCR with primers designed on the exon 2 of the *Ccr5* gene, we did not detect any band, indicating that the binding between NF-κB and *Ccr5* regulatory regions was specific (Figure 2A). In addition, pharmacological inhibition of NF-κB activation with the IkB-*α* phosphorylation inhibitor, BAY 11-7082, prevented the up-regulation of *Ccr5* expression in primary cortical neurons treated with H_2_O_2_ (Figure 2B). We also examined whether p65 activation after H_2_O_2_ exposure could be influenced by a CCR5 feedback mechanism, by examining the impact of *Ccr5* deficiency. Using confocal microscopy, we observed that the nuclear translocation of p65 after H_2_O_2_ exposure was similar in both CCR5^+/+^ and CCR5 knockout neurons (Figure 2C-D). Therefore, our results showed that in neurons i) NF-κB binding to *Ccr5* regulatory sequences elicited the expression of *Ccr5* and ii) oxidative stress induced NF-κB nuclear translocation that was not reinforced by a CCR5-dependent feedback loop.

**Figure 2.**
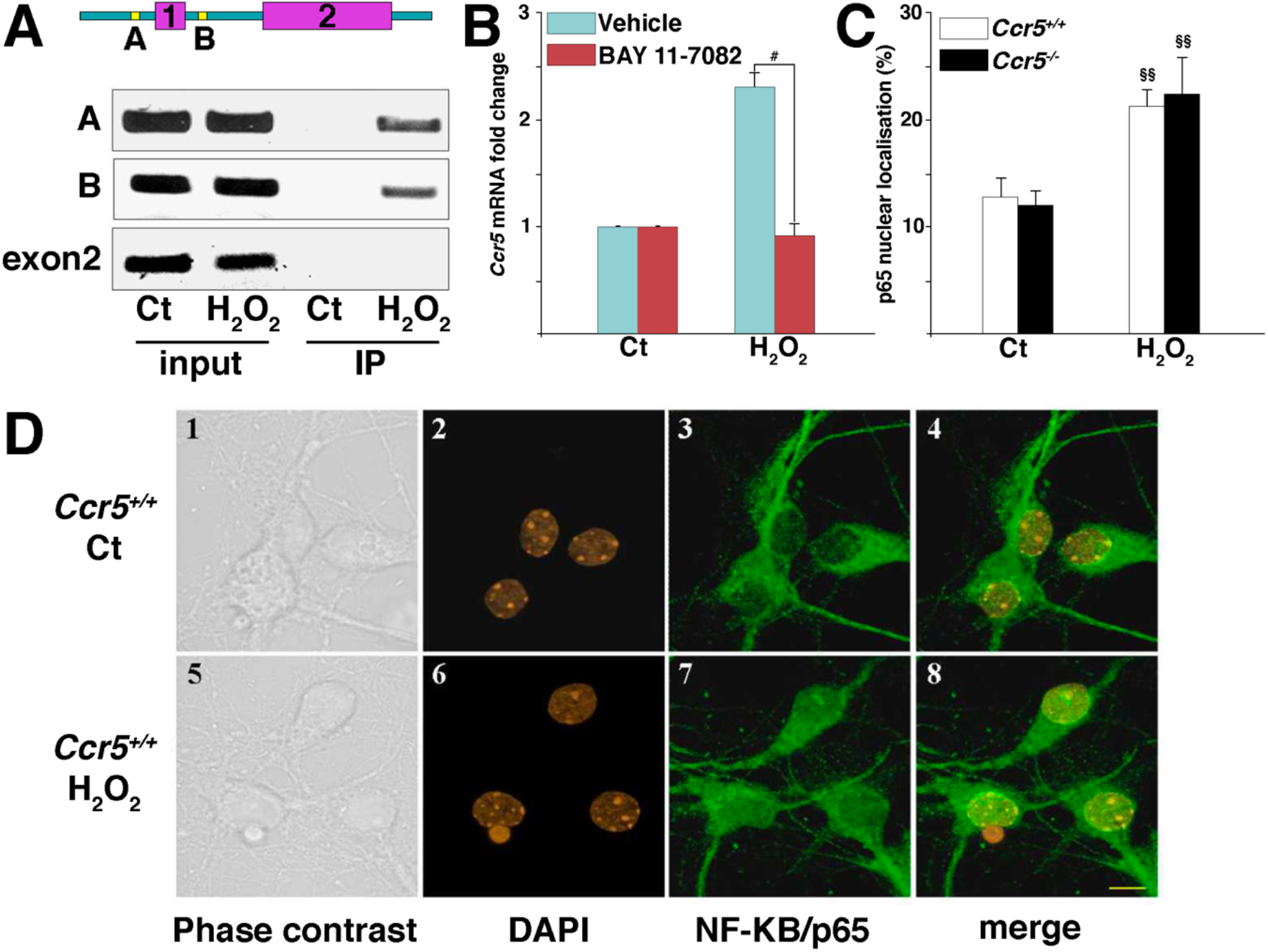
CCR5 expression in response to oxidative stress is mediated by NF-κB. **A**. *Upper*: Schematic overview of the murine *Ccr5* gene. Blue boxes represent introns, pink boxes exons, and yellow boxes (A and B) putative NF-κB/p65 binding elements. *Lower*: Chromatin immunoprecipitation using an NF-κB/p65 antibody of wild-type neurons after H_2_O_2_ (30 μM) or vehicle (Ct) treatment. The input samples represent equal fractions of DNA extract collected prior to immunoprecipitation. **B**. *Ccr5* mRNA levels measured by Real-Time PCR in wild-type neurons pretreated with the NF-κB inhibitor BAY 11-7082 and treated with H_2_O_2_ (30 μM). *#P*<0.05 using two-way ANOVA followed by Tukey *post-hoc* test (*n*=4). **C**. Nuclear localization of NF-κB/p65 staining in control (Ct) and H_2_O_2_-treated *Ccr5*^+/+^ and *Ccr5*^-/-^ primary neurons analyzed by an automated image program. ^§§^*p*<0.01 as compared to the respective control, using one-way ANOVA followed by Tukey *post-hoc* test. **D**. Confocal fluorescence microscopy showing cell morphology (phase contrast), nuclear counterstaining with DAPI (orange) and NF-κB/p65 staining (green). In the merged images, the nuclear localization of NF-κB is indicated in yellow (Scale bar=10 μm).

### Enhanced H_2_O_2_-elicited cell death in CCR5-deficient primary cortical neurons

We further investigated whether CCR5 has a functional role in the response to oxidative stress, and in particular in the regulation of neuronal survival. We therefore isolated primary cortical neurons from control and *Ccr5*-deficent mouse embryos, to assess their response to H_2_O_2_ independently of non-neuronal cells. After three hours of treatment with 30 μM H_2_O_2_, wild-type neurons did not exhibit major morphological alterations (Figure 3A) and only a modest decrease in cell number was observed (Figure 3B). In contrast, CCR5 knockout induced morphological alterations with visible signs of neurite degeneration in neurons exposed to H_2_O_2_ (Figure 3A). Also, the number of CCR5-deficient neurons was markedly diminished after H_2_O_2_ exposure (Figure 3B). Similar results were obtained with other quantitative cell viability assays by using Calcein-AM (Figure 3C) or AlamarBlue^®^ (Figure 3D). Although with different extents, due to the specific characteristics of each assay, in both cases we confirmed an increased death of primary neurons lacking CCR5 exposed to H_2_O_2_, as compared to wild type.

**Figure 3.**
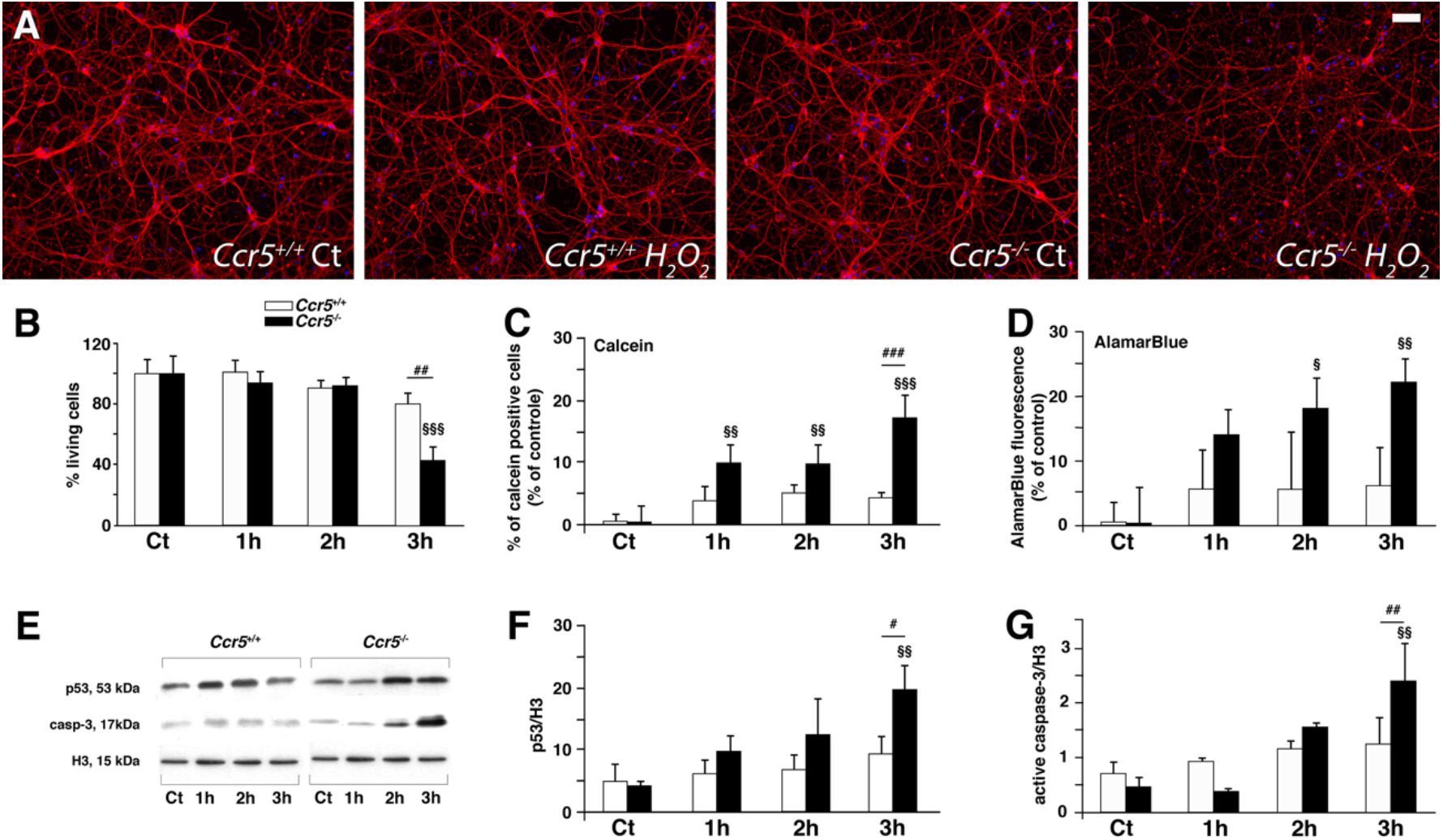
Oxidative stress induces cell death of Ccr5^-/-^ neurons. Primary cortical neurons at day 10 *in vitro*, were treated with H_2_O_2_ (30 μM) or vehicle (Ct) for 1-3 hours. **A**. Representative images of *Ccr5*^+/+^ and *Ccr5*^-/-^ primary neurons treated or not with H_2_O_2_. Scale bar: 50 μm. **B**. Percentage of living cells stained with *β*3-tubulin (red) and DAPI (blue) after H_2_O_2_ exposure. (**C-D**) percentage of dead cells after H_2_O_2_ exposure in wild-type and *Ccr5*^-/-^ neuronal cultures using Calcein (**C**) or AlamarBlue^®^ assay (**D**). **E**. Representative immunoblots of p53, cleaved caspase-3 (Casp-3) and histone H3 proteins in primary neurons treated with H_2_O_2_ (30 μM) for 1, 2 or 3 hours. (**F-G**) Optical density of p53 and cleaved caspase-3 bands normalized to the H3 protein values. *\*p*<0.05, *\*\*p*<0.01, using two-way ANOVA followed by Tukey *post-hoc* test (*n*=4). The Tukey post hoc tests indicate differences compared to control (§) and between to *Ccr5*^+/+^ and *Ccr5*^-/-^ primary neurons at the same treatment condition (#). The number of symbols indicates the significance level (1: *p*<0.05; 2: *p*<0.01; 3: *p*<0.001).

It has been demonstrated that p53 is one of the major mediators of ROS-induced apoptosis [22]. Therefore, we evaluated by Western blotting the levels of p53 and of the apoptotic marker caspase-3. In wild-type neurons, the levels of both proteins were not markedly affected by H_2_O_2_ exposure. In contrast, both p53 and active caspase-3 were significantly increased in a time-dependent manner in CCR5-deficient neurons (Figure 3E-G). Taken together, these data indicated that oxidative stress-induced cell death is increased in CCR5-deficient primary neurons and that CCR5 prevents oxidative stress-dependent activation of p53 and caspase 3.

## Discussion

Our study shows for the first time an association between the presence of the inactive human form CCR5-Δ32 (in combination with ApoEε4) and an increased risk of dementia, stronger for vascular and mixed dementia. The study in mice clarified the possible mechanisms leading to dementia. In fact, oxidative stress, a known actor during vascular damage, induces an increase in the neuronal expression of CCR5 and this absence in CCR5 deficient mice leads to the death of neurons. Thus, vascular damage inducing oxidative stress could lead to an increase in CCR5 in neurons which would have a protective role, while in its absence, neurons would be more vulnerable to apoptosis which could increase the risk of onset of vascular dementia. As a whole, we demonstrated that the human deletion of the *CCR5* gene (CCR5-Δ32 allele) acts in synergism with ApoEε4 as a major risk factor for dementia in elderly patients, specifically *CCR5* deletion increases the risk for vascular and mixed dementia, but not for AD.

The co-occurence of the ApoEε4/CCR5-Δ32 polymorphism combination shows a low frequency (44 / 1171 subjects from the combined Swiss and Italian cohorts) but is associated with a significant impact on the risk of vascular or mixed dementia. Importantly, this effect is observed even if 95% of CCR5 carriers are CCR5^+^/CCR5-Δ32 heterozygous. Thus, the subjects are not totally deficient in CCR5 (compared to the knockout mouse model) but the loss of a functional CCR5 allele is sufficient to increase the risk of dementia. The increased risk of vascular or mixed dementia is significant when considering both the original, and the pooled data set. However, the subjects of the Italian cohort showed a significant effect also of the ApoEε4/CCR5-Δ32 genotype for the AD group. This effect persisted in the analysis of the two cohorts together but with a low p value of 0.029. This difference between the two cohorts could at least partly be explained by a significantly younger age in the Italian cohort. In fact, when the analysis is carried out according to age (<80 and ≥80 year groups), the ApoEε4/CCR5-Δ32 polymorphism did not increase the risk for AD. This observation agrees with previous epidemiologic studies showing the absence of AD risk changes according to CCR5-Δ32 or the ApoEε4/CCR5-Δ32 combination [23-26]. These investigations showed differences with ours, especially in the experimental design. They used case control protocols, the mean age is sometime lower than herein (reducing the possibility of dementia) and they only included AD patients and controls. In our study, we prospectively included patients admitted to the Geneva University geriatric hospital in a randomized fashion. Thus, i) cognitively normal patients were chosen in a randomized manner and were considered as a control group; ii) the age of the patients included in the study is consistent with the distribution of dementia in the population of industrialized countries (average 85 years) and iii) all three major types of old age dementia were included in our patient population. An additional strength of this study is the fact that the same neuropsychologist carried out a systematic and complete neuropsychological assessment of all patients, increasing the accuracy of cognitive diagnosis. Thus, to our understanding our randomized patient collection methodology allows to study genetic factors in an authentic geriatric hospitalized population. Importantly, although CCR5-Δ32 alone did not have a significant impact on dementia in all group tested, the ApoEε4/CCR5-Δ32 combination increased the risk of dementia in both the Swiss and the Italian population, excepted when adjusted for age and sex in the Italian cohort. Even more important, although the risk of dementia is not modified by the ApoEε4/CCR5-Δ32 combination in subjects aged <80 years, it is drastically increased in subjects aged ≥80 years as compared to ApoEε4 alone (OR: 11.19 vs 2.42, respectively, including both Swiss and Italian populations). This specific approach has therefore made it possible to demonstrate that the ApoEε4/CCR5-Δ32 combination generates a higher risk for vascular or mixed dementia with a significant impact of age.

In order to hypothesize the mechanisms involved in the greater propensity of carriers of the knockdown form of CCR5 to vascular dementias, the second part of our study focused on the neuronal role of CCR5. In response to vascular damage, oxidative stress occurs and has been mimicked here by various stimuli. All of them induced neuronal expression of CCR5, thus extending to neurons the observations made on other cell types, in particular monocytes [27]. In addition, our data established redox-sensitive NF-κB activation as a mediator of *CCR5* expression in neurons. The demonstration of the translocation and then of the binding of NF-κB to the gene encoding CCR5 confirms the hypotheses made in the literature regarding the mode of activation of CCR5 [28-30]. To evaluate the role of such expression, the impact of oxidative stress was assessed in control and CCR5^-/-^ neurons. Morphological alterations with visible signs of neurite degeneration as well as neurochemical reorganisation with increased activated *caspase-3* and *p53* levels have been observed concomitantly with cell death. As both caspase-3 and p53 are involved in cell death by apoptosis [31], it can be concluded that the mechanism of H_2_O_2_-induced cell death is, at least in part, apoptosis. Neuronal death after nerve transection or ischemia are increased in CCR5-deficient mice corroborating the idea of an anti-apoptotic role of neuronal CCR5 [15, 16]. The neurotoxicity effect of the absence of CCR5 was previously hypothesized in the MPTP injection model of Parkinson’s disease, where the decrease in dopamine neurons was enhanced by CCR5 deficiency [32]. Brain damages in response to stroke are increased in CCR5 deficient mice, but, in contrast to such neuroprotective effect of CCR5, one study showed a better cognitive outcome in CCR5-Δ32 patients with stroke [33, 34]. A protective role of CCR5 was also observed in multiple sclerosis [35]. Thus, in response to oxidative stress the absence of neuronal activation of CCR5 mediated by NF-κB activation pathway confers neurovulnerability with induction of molecular players in apoptosis and cell death.

Additionally, to the neuroprotective role of neuronal CCR5, some studies have suggested that glial CCR5, and glial-neuron CCR5-dependent interactions result in cell survival and limitation of the release of inflammatory mediators [16, 36]. However, the CCR5 deletion does not appear to alter the recruitment of glial cells to the ischemic stroke site but could induce a neurotoxicity pattern at the microglia [15, 16, 37]. The kinetics of activation of the NF-κB pathway may also be of importance in the pro- or anti-apoptotic fate of the cellular response [38, 39]. In fact, a rapid activation elicits protective mechanisms, while a persistent activation induces the expression of pro-apoptotic factors. In our model, we observed that NF-κB is rapidly induced by H_2_O_2_ and determined the transactivation of the *Ccr5* gene, suggesting an anti-apoptotic effect on neurons. In contrast, in some other situations, CCR5 may (like NF-κB) have pro-apoptotic effects [40, 41]. Besides a role in neuronal response to oxidative stress, studies showing that the absence of CCR5 could play a role in the integrity of the blood vessels [42] also suggest the possibility of an implication in the appearance of vascular damage. This idea is reinforced by the increased ischemic risk in ApoEε4 carriers [43, 44]. Further studies are needed to better explore the full role of the CCR5-Δ32/ApoEε4 combination on the risk of cerebral vascular damages.

The potential clinical impact of our observations is on the level of risk prediction, as well as the development of novel treatment concepts. Hitherto, genetic risk factors in complex multigenetic diseases, such as old age dementia, do not contribute to patient diagnosis and management. However, the increase in risk of developing dementia with a vascular component for the combined ApoEε4/CCR5-Δ32 genotype is such that this notion might have to be reconsidered. Presently, vascular dementia is not curable but preventable [45]. Can the ApoEε4/CCR5-Δ32 constellation be used to predict conversion to dementia with a vascular component? In our study, only four cognitively normal ApoEε4/CCR5-Δ32 carriers were observed, thus the numbers are too small to obtain significant results regarding their conversion rate. All four subjects already converted to dementia with vascular component; three subjects within three years and one subject within four years since their inclusion in the study. Thus, the conversion rate of cognitively normal participants in our study as a function of the genotype was as follows: ApoEε4/CCR5-Δ32 100%, ApoEε4 alone 50%, CCR5-Δ32 alone 40%, none 30%. Thus, future studies will have to address the question whether the ApoEε4/CCR5-Δ32 genotype allows the identification of individuals presenting a higher risk to develop dementia with a vascular component and hence might benefit most from prevention measures. Finally, new treatments for vascular dementia might emerge from our observations into two different directions. First development could be tailored as a direct stimulation of CCR5 receptors in the central nervous system; second development could be based on a mechanistic understanding of neuroprotection by CCR5.

## Methods

### Patients

A prospective study was carried out at the Geneva University Hospitals, at the geriatrics hospital, Switzerland. Patients and data collection have been described in a previous study [46]. Briefly, patients were recruited by staff members with specific clinical training. The sampling frame consisted of a complete list of consecutive admissions of patients age range 65 - 99 year. A random sample was selected each day, using a computer-generated random table. The exclusion criteria were disorders interfering with psychometric assessment (severe deafness and blindness; major behavioural disturbances, such as severe aggressiveness, psychotic, suicidal behaviour, persistent delirium), terminal illness with an expected survival period of less than six weeks, and living outside of the Geneva area, due to difficulties in monitoring patients during follow-up. The local ethics committee approved the study protocol and signed written and informed consent was obtained from patients, their families or legal representatives. Patient history was recorded on a standardised form and a comprehensive assessment was performed by the same geriatrician (DZ). The methods to perform the cognitive diagnostic are given in Supplemental methods.

### Genotype analyses. CCR5 gene amplification and sequencing

The region of the CCR5 gene that flanks the 32 bp deletion was specifically amplified by PCR from genomic DNA with forward (TCCCAGGAATCATCTTTACCA) and reverse (AGGATTCCCGAGTAGCAGATG) primers and STAR DNA polymerase (Takara Bio Inc., Kyoto, Japan), according to the manufacturer’s instructions. Observed wild-type and deleted fragments were 183 bp and 151 bp, respectively. ***ApoE genotype***. The *ApoE* genotype was analysed similarly to *CCR5*, by sequencing PCR fragments obtained from the *ApoE* coding region (2795 to 3276) using specific primers. The sequence signals at positions 2901(T/C) and 3041(C/T) were read manually.

### Validation study

The validation study was carried out on DNA from a total of n=777 patients (n=319 AD, n=125 vascular dementia, n=176 mixed dementia) and from n=157 subjects with normal cognitive function (Table 3) age range 48 – 97 year. Clinical diagnosis for probable AD, vascular dementia and mixed dementia was made at the MAC Memory Clinic of the IRCCS Centro San Giovanni di Dio Fatebenefratelli (Brescia) according to international guidelines. DNA samples were available from the biological bank of IRCCS Fatebenefratelli Brescia, Italy. Written informed consent was obtained from all subjects.

### Isolation and culture of primary cortical neurons

Cortical neurons were prepared from CCR5^+/+^ and CCR5^-/-^ fetal brains at day E17.5 and were challenged with different chemicals. The detailed procedure is given in Supplemental methods.

### Real-Time quantitative and semi-quantitative end-point PCR

Real-time PCR (qPCR) reactions were performed using Power SYBR® Green PCR master mix (Applied Biosystems) and a Chromo 4TM Real-Time system (Bio-Rad). Quantification was performed at a threshold detection line (Ct value). The Ct value of each target genes was normalized against that of ribosomal protein S9 (*Rps9*) and TATA-box binding protein (*Tbp*) mRNAs used as housekeeping genes. The list of the primers used is given in Supplemental table 1.

### Immunofluorescence

Details of the procedure is provided in Supplemantal methods. Cells were then incubated overnight (4°C) with β3-tubulin (1:2000, Sigma-Aldrich) or NF-κB/p65 (1:500, Abcam) antibodies. Immunodetection was performed using Alexa 488 or Alexa 555 conjugated secondary antibodies (1:1000, Molecular Probes), followed by cell nucleus staining with a DAPI solution.

### Chromatin immunoprecipitation assay

Mouse primary cortical neurons were treated for 1 h with 30 μM H_2_O_2_ or vehicle. ChIP assay was then performed as previously described [47] and fully described in Supplemental methods. The immunoprecipitated DNA and the input chromatin were analyzed by end-point PCR (40 cycles) using promoter-specific primers (Supplemental table 1). The specificity of chromatin immunoprecipitation was assessed by PCR using primers located in the *Ccr5* exon 2.

### Calcein-AM and AlamarBlue^®^ assay

Primary neurons were seeded in 96-well plates and cultured as described above. After 10 DIV, neurons were exposed to 30 μM H_2_O_2_ (1h, 2h, 3h) or vehicle. Six replicates were performed for each condition. At the end of the treatment, medium was removed and a PBS solution with Calcein-AM (1:100, Invitrogen) or AlamarBlue^®^ (1:10, Invitrogen) was added to the cells for 40 minutes. Signals were read by using Fluostar Optima (BMG Labtech).

### Western blotting

Following chemical challenge, primary neurons were collected and lysed on ice (lysis buffer:50 mM Tris-HCl, pH 7.4, 50 mM NaCl, 10 mM MgCl_2_, 1 mM EGTA, 1% Triton X-100 supplemented with a protease inhibitor cocktail from Roche). A total of 30 μg of proteins per lane was diluted in loading buffer and denatured at 70°C for 10 minutes. Western blotting was performed with standard procedures using cleaved caspase-3 (1:500, R&D), p53 (1:500, Chemicon) and histone H3 (1:4000, Sigma-Aldrich) antibodies. Optical densities of the bands were measured using ImageJ software.

### Statistical methods

Continuous variables of human data are presented as means ± standard deviation (SD). Preclinical data are expressed as means ± standard error (SEM). Mann-Whitney u tests or Kruskal-Wallis ANOVA were used to compare data between cognitively normal and demented patients respectively between cognitively normal or patients affected with the main aetiologies of dementia. Univariate analysis was performed to identify independent risk factors associated with dementia in general and with the main aetiologies of dementia. Odds ratios (OR) and 95% confidence intervals (CI) were calculated. The variables assessed as possible predictors included age, sex, CCR5 and ApoEε4 gene polymorphisms the 3 latter being treated as binary variables. Multiple logistic regression analysis was then carried out to assess interactions between variables. Student’s *t*-test, one-way or two-way analysis of variance (ANOVA) followed by *post-hoc* Tukey test (as indicated in the figure legends) were used to analyse preclinical experiments. Statistical analyses were performed with Stata software version 14.1. For all tests, a *P* value inferior to 0.05 was taken as statistically significant.

## Data Availability

All data produced in the present study are available upon reasonable request to the authors

## Author contributions

DZ and K-HK design the research studies, search for the founding and collaborations, and analyzed the data. FH performed patients’ related statistical analysis. DZ included the patients, was responsible by the clinical and neuropsychological assessment and the clinical follow-up in Geneva. SS search for collaborations, conducted the translational experiments and analyzed the data. GB was responsible of the clinical and neuropsychological assessment in Brescia; RG and LB acquired genetic data. AM helped in basic experiments and review of the manuscript. BBT analyzed the data, integrated the translational data and wrote the manuscript. All the authors reviewed the manuscript.

## Acknowledgments

This work was supported by grants from the Swiss National Science Foundation (3200B0-102069 and 33CM30-124111), the Swiss Foundation for Ageing Research (AETAS, DZ) and by Italian Ministry of Health (Ricerca Corrente) (RG, LB, GB)

