## Supplemental methods for "CCR5 deficiency: decreased neuronal resilience to oxidative stress and increased risk of vascular dementia"

***Cognitive diagnostic*.**

To reduce the possibility of simultaneous delirium, a neuropsychologist assessed all included subjects one week after admission. The same neuropsychologist carried out a comprehensive complete standardised battery of neuropsychological tests [1]. Briefly, the neuropsychological testing included the Mini Mental Status Examination, the Mattis Dementia Rating Scale and the clinical dementia rating scale for global scores; the Buschke Double Memory Test and the Shapes Test to assess episodic memory; the digit span forward and backward and the Corsi Block-Tapping Test to assess working memory; the Boston Naming Test to assess language; The Verbal Fluency Test, the Trail Making Test and the clock-drawing Test to assess executive functions; The Digit Symbol Test for attention; the mental rotation task after Luria and the CERAD figures to measure spatial thinking abilities and the Stroop test to examine information-processing-time and interference-inhibition. Subsequently, the same geriatrician (DZ) applied the formal clinical criteria for the aetiology of dementia: DSM IV-TR for dementia [2], NINCDS-ADRDA [3] for Alzheimer's disease and NINDS-AIREN [4] for vascular and mixed dementia. Cerebral imaging was also used to support the subgroup classification of dementia for all patients. Patients were assigned to five groups: i) cognitively normal, ii) AD, iii) vascular dementia, iiii) mixed dementia and iiiii) other dementia.

***Isolation and culture of primary cortical neurons*.**

All experiments were approved by the ethical committee of the University of Geneva and the Cantonal Veterinary Office. After dissection, cerebral cortices were digested with Trypsin/EDTA (Gibco-Invitrogen) and dissociated mechanically. Primary neurons were cultured in neurobasal medium (Gibco-Invitrogen) supplemented with 2% B-27 Serum-Free supplement (Gibco-Invitrogen), 2 mM L-glutamine, 0.1 mM β-mercaptoethanol (Sigma-Aldrich) and 1% penicillin/streptomycin (Gibco-Invitrogen) at 37°C in 5% $CO_2$. At day 3 *in vitro*, they were treated with 5 μM cytosine β-D-arabinofuranoside (Sigma-Aldrich) to inhibit non-neuronal cell proliferation. Half of the medium was changed at day 5 *in vitro* and then every 3 days. Neuronal cultures were challenged by adding one of the following chemicals: hydrogen peroxide ($H_2O_2$, 30 μM; Sigma-Aldrich) glutamate (100 μM; TOCRIS Bioscience), or vehicles (PBS for $H_2O_2$, and ethanol for glutamate). Neurons treated with vehicles for the longer time period (3h) were considered as controls (ctr), as indicated in figure legends. Hypoxia was mimicked by adding cobalt chloride ($CoCl_2$, 500 μM, Calbiochem) [5] in a glucose-deprived medium. In this case, neurons incubated in the same medium supplied with glucose for 3h were considered as control. Incubation with the NF-κB inhibitor BAY 11-7085 (10 μM, Sigma-Aldrich) or vehicle (DMSO) was performed 30 min before adding $H_2O_2$, and maintained during the treatment with $H_2O_2$ or PBS (1h). All treatments were stopped at different time points by washing cultures with ice-cold PBS.

***Real-Time quantitative and semi-quantitative end-point PCR***.

Total RNA was isolated using RNeasy mini kit (Qiagen) according to the manufacturer's instructions. Residual genomic DNA was removed using RNase-Free DNase set (Qiagen). Total RNA (1 µg) was reverse transcribed using the superscript II kit according to the manufacturer's instructions (Invitrogen). Real-time PCR (qPCR) reactions were performed in triplicates for each condition. Semi-quantitative end-point PCR was performed by determining the suitable number of PCR cycles giving linear cDNA amplification of each gene of interest using Taq DNA polymerase (Qiagen) as previously described [6]. Amplification of the *Rpl32* housekeeping gene encoding the ribosomal protein L32 was used as control.

***Immunofluorescence***.

Neurons seeded on glass coverslip in 6-well plates were cultured as described above. After mild fixation (PBS/2% paraformaldehyde), cells were permeabilized (PBS/triton-x 0.1%) and non-specific binding sites were blocked with sera from animal species used to generate secondary antibodies. After incubation with primary and secondary antibodies, and DAPI, the neuronal survival was determined by analyzing nucleus morphology (pyknotic profile corresponding to chromatin condensation) of cells immunoreactive for β3-tubulin. For each experiment, cells were counted blindly in five different fields, randomly selected, and the percentage of pyknotic cells was determined ($n$=4). Confocal imaging was performed as described above. Confocal images were analyzed using the Metamorph software (Molecular Devices) and the overlapping of DAPI and NF-κB subunit p65 signals was quantified blindly in cells from five different fields per experiment, randomly selected ($n$=4).

***Chromatin immunoprecipitation assay.***

Cross-linking was performed by incubating cells for 8 min at room temperature with 5 mM HEPES, pH 7.9, 10 mM NaCl, and 1.1% formaldehyde. Cross-linking was stopped by adding 180 mM glycine. After one wash with PBS, cells were lysed by pipetting up and down in ice-cold 10 mM Tris-HCl, pH 8.0, 0.5% NP-40 supplemented with protease inhibitor cocktail (Roche). Nuclei were pelleted and lysed in 10 mM Tris-HCl, pH 8.0, 500 mM NaCl, 1% Triton X-100, 0.5% sodium deoxycholate supplemented with protease inhibitor cocktail. Cross-linked chromatin was sheared into 600-200 base pairs fragments by sonication and cleared by centrifugation at 15,000 *g* for 15 min. A total of 20 µg of chromatin supernatants was diluted (1:10) in ChIP buffer [200 mM Hepes, pH 7.9, 2 M NaCl, 20 mM EDTA] supplemented with 200 µg/mL salmon-sperm DNA and protease inhibitor cocktail. After preclearing the supernatants by incubation for 30 min at 4°C with protein A–sepharose beads (Pierce), half of the chromatin was stored at -20°C and used as control chromatin input. The rest of the supernatant was then incubated overnight at 4°C with 2 µg of NF-κB subunit p65 antibody (Abcam), cleared by centrifugation at room temperature for 10 min at 8,000 *g* before 2 h of incubation at room temperature in 20 µL protein A–sepharose beads. The beads were washed as follows: twice in ChIP buffer; twice in ChIP buffer supplemented with 300 mM NaCl, 1% Triton X-100, 0.1% sodium deoxycholate; twice in Tris-HCl, pH 8.0, 250 mM LiCl, 2 mM EDTA, 0.5% sodium deoxycholate; once in 10 mM Tris-HCl, pH 8.0, 1 mM EDTA, 0.1% NP-40. Immune complexes were eluted by incubating 10 min at 65°C in 111 mM Tris-HCl, pH 8.0, 1.11% SDS. Cross-links of the immunoprecipitated DNA and of the chromatin input were reversed to detach DNA from proteins by incubation for 2 h at 42°C after addition of 100 mM NaCl containing 20 mg/ml Proteinase K (Qiagen) and further incubation at 67°C overnight. After extraction with phenol-chloroform-isoamyl alcohol (25:24:1) and chloroform-isoamyl alcohol (24:1), DNA was precipitated with 100% ethanol in the presence of 20 µg glycogen and 0.3 M sodium acetate. Chromatin pellets were resuspended in 100 µL TE buffer.


[1] Adler MW, Geller EB, Chen X, Rogers TJ (2006) Viewing chemokines as a third major system of communication in the brain. *AAPS J* **7**, E865-870.

[2] Association AP (2000) Diagnostic and Statistical Manual of Mental Disorders, 4th Ed Text Revision (DSM IV-TR). *Washington, DC: American Psychiatric Association*.

[3] McKhann G, Drachman D, Folstein M, Katzman R, Price D, Stadlan EM (1984) Clinical diagnosis of Alzheimer's disease: report of the NINCDS-ADRDA Work Group under the auspices of Department of Health and Human Services Task Force on Alzheimer's Disease. *Neurology* **34**, 939-944.

[4] Roman GC, Tatemichi TK, Erkinjuntti T, Cummings JL, Masdeu JC, Garcia JH, Amaducci L, Orgogozo JM, Brun A, Hofman A, et al. (1993) Vascular dementia: diagnostic criteria for research studies. Report of the NINDS-AIREN International Workshop. *Neurology* **43**, 250-260.

[5] Rossignol F, de Laplanche E, Mounier R, Bonnefont J, Cayre A, Godinot C, Simonnet H, Clottes E (2004) Natural antisense transcripts of HIF-1alpha are conserved in rodents. *Gene* **339**, 121-130.

[6] Bonnefont J, Daulhac L, Etienne M, Chapuy E, Mallet C, Ouchchane L, Deval C, Courade JP, Ferrara M, Eschalier A, Clottes E (2007) Acetaminophen recruits spinal p42/p44 MAPKs and GH/IGF-1 receptors to produce analgesia via the serotonergic system. *Mol Pharmacol* **71**, 407-415.
