## Supplemental for "CCR5 deficiency: decreased neuronal resilience to oxidative stress and increased risk of vascular dementia"

**Supplemental data**

| Gene | Primer sequences | Product size | Tm | # cycles |
|---|---|---|---|---|
| ***Ccr5* (qPCR)** | F: GTT CCT GAA AGC GGC TGT AAA<br>R: GCA GTC AGG CAC ATC CAT AGA C | 74 | - | - |
| ***Cxcr4* (qPCR)** | F: TGG CAT AGT CGG CAA TGG A<br>R: CGT CAT GCT CCT TAG CTT CTT CT | 66 | - | - |
| ***Tbp* (qPCR)** | F: TTGACCTAAAGACCATTGCACTTC<br>R: TTCTCATGATGACTGCAGCAAA | 78 | - | - |
| ***Rps9* (qPCR)** | F: GAC CAG GAG CTA AAG TTG ATT GGA<br>R: TCT TGG CCA GGG TAA ACT TGA | 81 | - | - |
| ***Ccl3*** | F: ATG AAG GTC TCC ACC ACT GC<br>R: CCC AGG TCT CTT TGG AGT CA | 279 | 60 | 32 |
| ***Ccl4*** | F: GCC CTC TCT CTC CTC TTG CT<br>R: GTC TGC CTC TTT TGG TCA GG | 196 | 60 | 32 |
| ***Ccl5*** | F: CCC TCA CCA TCA TCC TCA CT<br>R: CCT TCG AGT GAC AAA CAC GA | 185 | 60 | 32 |
| ***L32*** | F: GTG AAG CCC AAG ATC GTC AA<br>R: TTG GTG ACT CTG ATG GCC AG | 349 | 58 | 26 |
| ***Ccr5 p*romoter site A** | F: CTT GAC TCA AAT GTG GGC TTT<br>R: GGC TGA GAG GTG ACT TAC CA | 400 | 58 | 40 |
| ***Ccr5* promoter site B** | F: TTT AAA CAG GGC AAG CCA GT<br>R: TTC TCA GCT GCA AGA AGC AA | 379 | 58 | 40 |

**Supplemental table 1.** Oligonucleotide primers and conditions used for real-time PCR (qPCR) and semi-quantitative end-point RT-PCR.Ccl: Chemokine (C-C motif) ligand; Ccr5: C-C chemokine receptor type 5; Rps9: Ribosomal Protein S9; Tbp: TATA-binding protein

| | Cognitively normal subjects | | All type of dementia | | Alzheimer's disease | | Vascular and mixed dementia[A] | |
|---|---|---|---|---|---|---|---|---|
| **Age[B,C]** | 80.3 | 7.8 | 81.1 | 7.0 | 81.8 | 6.1 | 80.3 | 7.6 |
| **Female[C]** | 238 | 67.7% | 592 | 73.2% | 306 | 78.1% | 276 | 68.5% |
| **ApoEε4$^-$ / CCR5$^+$** | 261 | 72.1% | 414 | 51.2% | 197 | 50.3% | 207 | 51.4% |
| **ApoEε4$^-$ / CCR5-Δ32** | 49 | 13.5% | 86 | 10.6% | 41 | 10.5% | 41 | 10.2 |
| **ApoEε4$^+$ / CCR5$^+$** | 46 | 12.7% | 271 | 33.5% | 138 | 35.2% | 133 | 33% |
| **ApoEε4$^+$ / CCR5-Δ32** | 6 | 1.7% | 38 | 4.7% | 16 | 4.1% | 22 | 5.5% |
| **Total** | 362 | 100.0% | 809 | 100.0% | 392 | 100.0% | 403 | 100.0% |

**Supplemental table 2.** A comparison of CCR5-Δ32 and ApoEε4 allele frequencies between dementia patients, including the various dementia aetiologies, and subjects that are cognitively normal in the Italian and Swiss samples. To measure the impact of cerebral vascular lesions, patients with vascular and mixed dementia were analysed in the same group. Data are expressed as number of cases and %. [a]Data are expressed as means ± SD. ApoEε4+: one or two copies of ApoEε4; ApoEε4-: no copies of ApoEε4; CCR5Δ32+: one or two copies of the CCR5-32 bp deleted allele; CCR5Δ32-: two copies of the CCR5 wild-type allele.

| | *Dementia* (n = 809) | | | *Alzheimer's disease* (n = 392) | | | *Vascular or mixed dementia* (n = 403) | | |
|---|---|---|---|---|---|---|---|---|---|
| | **Univariate logistic regression** | | | | | | | | |
| | Crude OR | 95% CI | p | Crude OR | 95% CI | p | Crude OR | 95% CI | p |
| ApoEε4+A | **3.08C** | **2.20-4.31** | **<0.001** | **3.21** | **2.20-4.69** | **<0.001** | **3.25** | **2.25-4.71** | **<0.001** |
| Italy vs Swiss | **3.80** | **2.90-4.97** | **<0.001** | **5.11** | **3.65-7.15** | **<0.001** | **3.48** | **2.54-4.77** | **<0.001** |
| CCR5-Δ32B | 1.37 | 0.95-1.99 | 0.095 | 1.22 | 0.78-1.91 | 0.374 | 1.49 | 0.97-2.28 | 0.068 |
| Italy vs Swiss | **4.44** | **3.40.5.81** | **<0.001** | **5.80** | **7.17-8.08** | **<0.001** | **4.08** | **2.98-5.59** | **<0.001** |
| Age | **1.08** | **1.06-1.11** | **<0.001** | **1.14** | **1.11-1.18** | **<0.001** | **1.07** | **1.04-1.10** | **<0.001** |
| Italy vs Swiss | **7.84** | **5.64-10.9** | **<0.001** | **17.41** | **10.9-27.8** | **<0.001** | **6.98** | **4.71-10.3** | **<0.001** |
| Male vs female | **0.63** | **0.48-0.84** | **<0.001** | **0.42** | **0.29-0.60** | **<0.001** | 0.79 | 0.57-1.08 | 0.141 |
| Italy vs Swiss | **4.41** | **3.38-5.76** | **<0.001** | **6.37** | **4.53-8.94** | **<0.001** | **3.93** | **2.89-5.35** | **<0.001** |
| | **Multiple logistic regression** | | | | | | | | |
| | Adjusted OR | 95% CI | p | Adjusted OR | 95% CI | p | Adjusted OR | 95% CI | p |
| *Association ApoEε4 / CCR5Δ32* | | | | | | | | | |
| ApoEε4- / CCR5+ | 1.00 | -- | -- | 1.00 | -- | -- | 1.00 | -- | -- |
| ApoEε4- / CCR5-Δ32 | 1.44 | 0.95-2.16 | 0.083 | 1.39 | 0.85-2.29 | 0.191 | 1.46 | 0.90-2.38 | 0.123 |
| ApoEε4+ / CCR5+ | **3.11** | **2.17-4.46** | **<0.001** | **3.35** | **2.23-5.02** | **<0.001** | **3.18** | **2.14-4.73** | **<0.001** |
| ApoEε4+ /CCR5-Δ32 | **4.51** | **1.83-11.1** | **0.001** | **3.68** | **1.32-10.3** | **0.013** | **5.76** | **2.21-15.0** | **<0.001** |
| Italy vs Swiss | **3.95** | **3.00-5.20** | **<0.001** | **5.23** | **3.72-7.34** | **<0.001** | **3.70** | **2.68-5.12** | **<0.001** |
| *Adjusted for age and sex* | | | | | | | | | |
| ApoEε4- / CCR5+ | 1.00 | -- | -- | 1.00 | -- | -- | 1.00 | -- | -- |
| ApoEε4- / CCR5Δ32 | 1.42 | 0.93-2.17 | 0.108 | 1.25 | 0.73-2.15 | 0.414 | 1.50 | 0.91-2.48 | 0.109 |
| ApoEε4+ / CCR5+ | **3.21** | **2.22-4.63** | **<0.001** | **3.70** | **2.39-5.72** | **<0.001** | **3.22** | **2.15-4.82** | **<0.001** |
| ApoEε4+ /CCR5Δ32 | **4.71** | **1.82-12.2** | **0.001** | **4.03** | **1.15-14.1** | **0.029** | **5.94** | **2.19-16.1** | **<0.001** |
| Age | **1.08** | **1.06-1.10** | **<0.001** | **1.14** | **1.10-1.18** | **<0.001** | **1.07** | **1.04-1.10** | **<0.001** |
| Male vs female | **0.69** | **0.51-0.93** | **0.014** | **0.42** | **0.28-0.62** | **<0.001** | 0.83 | 0.59-1.16 | 0.28 |
| Italy vs Swiss | **7.29** | **5.18-10.3** | **<0.001** | **16.67** | **10.3-27.0** | **<0.001** | **6.78** | **4.49-10.2** | **<0.001** |

**Supplemental Table 3.** Univariate and multiple logistic regression models of the risk of dementia and of its main aetiologies with cognitively normal subjects as the reference group (n = 362) according to CCR5 and ApoE genotypes in the Italian and Swiss population. Models are adjusted by country.

[A]ApoEε4+: one or two copies of ApoEε4; ApoEε4-: no copies of ApoEε4; [B]CCR5Δ32+: one or two copies of the CCR5-32 bp deleted allele; CCR5Δ32-: two copies of the CCR5 wild-type allele. [C]Bold entries = relevant results.

| | *Dementia* (n = 317) | | | *Alzheimer's disease* (n = 126) | | | *Vascular or mixed dementia* (n = 186) | | |
|---|---|---|---|---|---|---|---|---|---|
| | | | | **Univariate logistic regression** | | | | | |
| | Crude OR | 95% CI | p | Crude OR | 95% CI | p | Crude OR | 95% CI | p |
| ApoEε4+A | **4.19C** | **2.60-6.76** | **<0.001** | **4.46** | **2.56-7-77** | **<0.001** | **4.17** | **2.49-6.99** | **<0.001** |
| Italy vs Swiss | **2.87** | **1.70-4.86** | **<0.001** | **3.81** | **1.75-8.30** | **0.001** | **3.34** | **1.75-6.36** | **<0.001** |
| CCR5-Δ32B | 0.95 | 0.53-1.69 | 0.859 | 0.77 | 0.35-1.69 | 0.509 | 1.13 | 0.59-2.18 | 0.706 |
| Italy vs Swiss | **3.66** | **2.18-6.14** | **<0.001** | **4.59** | **2.13-9.90** | **<0.001** | **4.36** | **2.30-8.29** | **<0.001** |
| Age | **1.05** | **1.00-1.10** | **0.036** | **1.18** | **1.08-1.28** | **<0.001** | 1.03 | 0.98-1.08 | 0.248 |
| Italy vs Swiss | **3.96** | **2.37-6.62** | **<0.001** | **6.61** | **2.96.14.77** | **<0.001** | **4.45** | **2.37-8.36** | **<0.001** |
| Male vs female | 0.94 | 0.64-1.39 | 0.759 | 0.67 | 0.40-1.10 | 0.114 | 1.05 | 0.68-1.63 | 0.828 |
| Italy vs Swiss | **3.70** | **2.23-6.14** | **<0.001** | **5.18** | **2.42-11.1** | **<0.001** | **4.24** | **2.27-7.91** | **<0.001** |
| | | | | **Multiple logistic regression** | | | | | |
| | Adjusted OR | 95% CI | p | Adjusted OR | 95% CI | p | Adjusted OR | 95% CI | p |
| *Association ApoEε4 / CCR5Δ32* | | | | | | | | | |
| ApoEε4- / CCR5+ | 1.00 | -- | -- | 1.00 | -- | -- | 1.00 | -- | -- |
| ApoEε4- / CCR5-Δ32 | 1.14 | 0.59-2.18 | 0.699 | 1.00 | 0.40-2.45 | 0.994 | 1.32 | 0.62-2.80 | 0.471 |
| ApoEε4+ / CCR5+ | **4.53** | **2.71-7.59** | **<0.001** | **4.80** | **2.66-8.66** | **<0.001** | **4.47** | **2.57-7.79** | **<0.001** |
| ApoEε4+ /CCR5-Δ32 | 2.72 | 0.82-9.05 | 0.103 | 2.22 | 0.44-11.14 | 0.331 | 3.43 | 0.95-12.4 | 0.059 |
| Italy vs Swiss | **2.86** | **1.67-4.89** | **<0.001** | **3.57** | **1.61-7.90** | **0.002** | **3.45** | **1.78-6.70** | **<0.001** |
| *Adjusted for age and sex* | | | | | | | | | |
| ApoEε4- / CCR5+ | 1.00 | -- | -- | 1.00 | -- | -- | 1.00 | -- | -- |
| ApoEε4- / CCR5Δ32 | 1.09 | 0.57-2.11 | 0.788 | 1.01 | 0.40-2.56 | 0.980 | 1.31 | 0.61-2.79 | 0.488 |
| ApoEε4+ / CCR5+ | **4.39** | **2.61-7.37** | **<0.001** | **4.44** | **2.42-8.16** | **<0.001** | **4.42** | **2.53-7-75** | **<0.001** |
| ApoEε4+ /CCR5Δ32 | 2.65 | 0.79-8-93 | 0.116 | 2.36 | 0.41-13.47 | 0.334 | 3.40 | 0.94-12.3 | 0.063 |
| Age | 1.03 | 0.98-1.08 | 0.207 | **1.14** | **1.04-1.24** | **0.003** | 1.01 | 0.96-1.06 | 0.764 |
| Male vs female | 0.90 | 0.60-1.37 | 0.630 | 0.61 | 0.35-1.07 | 0.084 | 0.99 | 0.62-1.58 | 0.964 |
| Italy vs Swiss | **2.99** | **1.74-5.15** | **<0.001** | **4.96** | **2.14-11.52** | **<0.001** | **3.49** | **1.79-6.82** | **<0.001** |

**Supplemental Table 4.** Bivariate and multiple logistic regression models of the risk of dementia and of its main aetiologies with cognitively normal subjects as the reference group (n = 177) according to CCR5 and ApoE genotypes in the Italian and Swiss populations aged < 80. Models are adjusted by country.

| | *Dementia* (n = 189) | | | *Alzheimer's disease* (n = 73) | | | *Vascular or mixed dementia* (n = 102) | | |
|---|---|---|---|---|---|---|---|---|---|
| | **Univariate logistic regression** | | | | | | | | |
| | Crude OR | 95% CI | p | Crude OR | 95% CI | p | Crude OR | 95% CI | p |
| ApoEε4$^{+}$[A] | **2.51**[C] | **1.53-4.12** | **<0.001** | **2.63** | **1.48-4.69** | **0.001** | **2.75** | **1.58-4.78** | **<0.001** |
| Italy vs Swiss | **10.88** | **6.95-17.0** | **<0.001** | **16.44** | **10.01-27.0** | **<0.001** | **8.12** | **4.96-13.3** | **<0.001** |
| CCR5-Δ32[B] | **1.70** | **1.02-2.84** | **0.041** | 1.34 | 0.71-2.51 | 0.370 | **1.98** | **1.10-3.56** | **0.022** |
| Italy vs Swiss | **12.16** | **7.77-19.0** | **0.000** | **17.85** | **10.9-29.2** | **<0.001** | **9.19** | **5.59-15.1** | **<0.001** |
| Age | 1.04 | 0.99-1.09 | 0.099 | 1.04 | 0.99-1.10 | 0.135 | 1.05 | 1.00-1.11 | 0.056 |
| Italy vs Swiss | **13.37** | **8.30-21.5** | **<0.001** | **20.85** | **12.1-36.0** | **<0.001** | **10.26** | **6.00-17.5** | **<0.001** |
| Male vs female | **0.53** | **0.34-0.82** | **0.005** | **0.33** | **0.18-0.59** | **<0.001** | 0.63 | 0.38-1.04 | 0.072 |
| Italy vs Swiss | **11.85** | **7.57-18.5** | **<0.001** | **19.73** | **11.7-33.1** | **<0.001** | **8.66** | **5.30-14.1** | **<0.001** |
| | **Multiple logistic regression** | | | | | | | | |
| | Adjusted OR | 95% CI | p | Adjusted OR | 95% CI | p | Adjusted OR | 95% CI | p |
| *Association ApoEε4 / CCR5Δ32* | | | | | | | | | |
| ApoEε4$^{-}$ / CCR5$^{+}$ | 1.00 | -- | -- | 1.00 | -- | -- | 1.00 | -- | -- |
| ApoEε4$^{-}$ / CCR5-Δ32 | 1.62 | 0.92-2.82 | 0.093 | 1.41 | 0.70-2.82 | 0.336 | 1.72 | 0.89-3.33 | 0.106 |
| ApoEε4$^{+}$ / CCR5$^{+}$ | **2.35** | **1.38-4.01** | **0.002** | **2.63** | **1.42-4.87** | **0.002** | **2.42** | **1.32-4.42** | **0.004** |
| ApoEε4$^{+}$ /CCR5-Δ32 | **7.65** | **1.70-34.5** | **0.008** | 4.86 | 0.88-26.8 | 0.07 | **11.19** | **2.36-53.0** | **0.002** |
| Italy vs Swiss | **11.40** | **7.26-17.9** | **<0.001** | **16.6** | **10.08-27.3** | **<0.001** | **8.91** | **5.39-14.7** | **<0.001** |
| *Adjusted for age and sex* | | | | | | | | | |
| ApoEε4$^{-}$ / CCR5$^{+}$ | 1.00 | -- | -- | 1.00 | -- | -- | 1.00 | -- | -- |
| ApoEε4$^{-}$ / CCR5Δ32 | 1.64 | 0.93-2.90 | 0.086 | 1.39 | 0.68-2.81 | 0.364 | 1.76 | 0.90-3.43 | 0.098 |
| ApoEε4$^{+}$ / CCR5$^{+}$ | **2.53** | **1.48-4.34** | **0.001** | **3.15** | **1.65-6.00** | **<0.001** | **2.57** | **1.40-4.74** | **0.002** |
| ApoEε4$^{+}$ /CCR5Δ32 | **7.48** | **1.64-34.1** | **0.009** | 6.11 | 0.94-39.6 | 0.058 | **10.55** | **2.21-50.2** | **0.003** |
| Age | 1.05 | 1.00-1.10 | 0.056 | 1.05 | 0.99-1.11 | 0.1 | **1.06** | **1.00-1.12** | **0.038** |
| Male vs female | 0.51 | 0.33-0.81 | 0.004 | 0.29 | 0.16-0.53 | 0.000 | 0.64 | 0.38-1.08 | 0.093 |
| Italy vs Swiss | **13.73** | **8.38-22.5** | **<0.001** | **22.68** | **12.6-40.8** | **<0.001** | **11.60** | **6.58-20.4** | **<0.001** |

**Supplemental Table 5.** Bivariate and multiple logistic regression models of the risk of dementia and of its main aetiologies with cognitively normal subjects as the reference group (n = 185) according to CCR5 and ApoE genotypes in the Italian and Swiss populations aged ≥ 80. Models are adjusted by country.
